# Rapid Support for Older Adults during the initial stages of the COVID-19 Pandemic: Results from a Geriatric Psychiatry helpline

**DOI:** 10.1101/2020.10.29.20218750

**Authors:** Anna-Sophia Wahl, Gloria S. Benson, Lucrezia Hausner, Sandra Schmitt, Annika Knoll, Adriana Feretti Bondy, Dimitri Hefter, Lutz Frölich

## Abstract

**Background:** The COVID-19 pandemic and governmental lockdown measures disproportionally impacts older adults. This study presents the results from a psychiatric helpline for older adults in Mannheim, Germany, during the lockdown, set up to provide information and psychosocial support measures. We aim to elucidate the needs of older adults, their reported changes and the psychological impact during the initial stages of the health crisis.

**Methods:** 55 older adults called the psychiatric helpline between April and June 2020. Information on demographics, previous diagnosis of psychiatric and somatic diseases as well as changes in daily life due to the pandemic was collected anonymously. Current mental health status was assessed using the depression HAMD-7 and the anxiety HAM-A scales.

**Results:** Most callers were women, older adults (*M* = 74.69 years), single and retired. 69% of callers reported new or an increase in psychiatric symptoms, with anxiety and depressive symptoms being the most common ones. Age was significantly negatively correlated to higher levels of anxiety and depression symptoms. Individuals with a previous diagnosis of a psychiatric disease reported significantly higher levels of depressive and anxiety symptoms than those without a diagnosis.

**Conclusion:** In older adults, the perceived psychological impact of the COVID-19 crisis appears to ameliorate with age. Individuals with a history of psychiatric disease are most vulnerable to negative mental health outcomes. Rapid response in the form of a geriatric helpline are useful initiatives to support the needs and the psychological well-being of older adults during a health crisis.

## Introduction

The COVID-19 pandemic has caused an unpreceded disruption of daily life around the world. Older adults over the age of 65 are considered being at higher risk of severe complications and mortality from COVID-19, and thus are disproportionally impacted (Grasselli et al., 2020; Jordan et al., 2020; Wang et al., 2020; Zhou et al., 2020). Worldwide governmental virus-containment-efforts include a variety of measures from social distancing to stay-at-home orders. The disruption of daily life through new imposed measures that increase isolation may have a stronger impact in older adults, which could in turn have deleterious effects on their mental health and well-being. Research focusing on the implications and effects of the COVID-19 pandemic and efforts that address the needs of older adults remain very limited.

Evidence is rising that mental health effects of the political measures for social distancing and isolation in older adults should not be neglected (Xiang et al., 2020). Previous literature shows that social isolation in older adults has been associated with increased depression and suicidality as well as psychoendocrine, metabolic and decreased immunological response (Steven W. Cole et al., 2015; Steve W. Cole et al., 2007; Santini et al., 2020). Several recent studies from different countries investigated the psychological impact of COVID-10 on mental health, (Rodriguez-Rey et al, Benke et al, 2020, (Luo et al., 2020) with a few focusing specifically in older adults ((Davies, 2020), (Meng et al., 2020). Their findings suggest that older adults experienced depression and anxiety, with older women experiencing more symptoms than older men (Meng et al., 2020). Social impact and attitudes also differ among older adults. A majority of older adults were worried about the future, feeling stress and anxiety. Younger older adults in their 60s were least optimistic the outcome and the duration of the pandemic than those aged 70 and older (Davies, 2020). Conversely, a meta-analysis using data from 27 countries found that older adults are not more willing to stricter adhere to new rules of social distancing and self-protection than other age groups (Daoust, 2020).

As daily infections increased globally in March, Germany started to implemented public health regulations. From March 22 until April 19, 2020 German states implemented “lock-down” restriction measures (e.g. stay-at home orders, closure of non-essential shops, minimum distance of 1.5 m to others, restriction to visits clinics and nursing homes and decreased social contact). During this initial wave of the COVID-19 pandemic in Germany we initiated a geriatric helpline to address and ameliorate the negative impact of the health crisis among older adults. The aim was to 1) provide information about COVID-19 and protective measurements, 2) issue practical help in case of problems with daily living and 3) provide medical expertise and psychological support for psychiatric symptoms. Furthermore, we established the helpline to assess the mental health impact in the aging population in real time, in order to minimize and ameliorate the adverse impact. In this ongoing project, we have analyzed a first cohort of participants from this helpline, to better understand the effect of the pandemic and of local lockdown measures on mental health-related issues in older adults. We provide demographic data, the impact of daily living and social interactions, as well as associations between sociodemographic variables and mental health status

## Methods

### Establishment of a Covid-19 hotline

3 weeks after the first enforced shut-down measurements in Germany, the geriatric helpline went live on April 13^th^, we established a COVID-19 helpline for older adults (>65 years old) at the Central Institute of Mental Health, Mannheim (CIMH), University of Heidelberg. The CIMH is the only psychiatric institution in the city of Mannheim, and provides full sectorized care for older adults (20% of 300,000 inhabitants). The goal of the hotline was to provide (1) information about the Covid-19 virus and education of meaningful rules of conduct, (2) psychiatric and psychological counseling, (3) acute psychotherapeutic interventions, (4) psychosocial support for a better daily structure of during the quarantine, (5) information of local care providers for assistance with daily living activities. Before the establishment of the helpline, information on local care providers for older adults (e.g. support on groceries shopping and other errands) from different organizations (governmental and non-governmental) was collected. Three psychiatrists and three psychologists from the gerontopsychiatry department and the memory clinic (with specialization in diagnosis and treatment of geriatric depression, anxiety and dementia) hosted the hotline. The hotline was announced in several local newspapers, radio stations as well advertised on the homepage of the city of Mannheim.

Callers could anonymously tell their needs and received advice, and calls were not recorded. However, callers were initially told that the information provided would be documented anonymously and further analyzed in a scientific manner. Verbal consent was acquired. We received 53 calls within a radius of 50-70km around Mannheim and two calls a longer way off (>100km).

A paper-based survey was developed to assess demographic information, previous diagnosis of psychiatric diseases, current psychiatric treatment or psychotherapy experience, somatic diseases, number of medications taken, new or increase of psychiatric symptoms, number of daily social interactions before the pandemic and during the pandemic, living and financial situation and requirements of daily support by others. The Charlson comorbidity index (CCI) was used to assess the age-comorbidity of somatic disease (Charlson et al., 1994). Mental health status was systematically assessed using the Hamilton depression and Hamilton anxiety scores.

### Mental Health Status

#### Depression

The 7-Item Hamilton Depression Rating Scale (HAMD-7) (McIntyre et al., 2002) is commonly used in clinical and research setting to evaluate the presence and severity of depression symptoms. Seven items are assessed in a semi-structure interview to asses: depressed mood, guilt, anhedonia, psychic anxiety, loss of energy (fatigue), somatic anxiety and suicidal ideation. The cutoffs for the different levels of depressive symptoms were set according to McIntyre: Scoring below 4 indicated no depressive symptoms (level I). A score of 4-11 was considered as mild (level II), a score of 12-20 as moderate (level III) and a score >20 as severe depression (level IV).

#### Anxiety

The Hamilton Anxiety Rating Scale (HAM-A) 14-Item version was used to asses and quantify the presence and severity of anxiety symptoms (Hamilton, 1959). The commonly used 14-Items version, as a clinician-rated semi-structured interview was used. Higher scores indicated greater anxiety symptoms severity (Katherine Shear et al., 2001). According to (Hamilton, 1959) a score of <17 indicates none to mild levels of anxiety (level I), a score of 18-24 moderate (level II), a score of 25-30 moderate to severe (level III) and a score >30 severe levels (level IV) of anxiety.

#### Data analysis

Data was entered into electronic records for further analysis. Statistical analysis was performed with SPSS Version 24. The descriptive statistics of the sample were computed for the sociodemographics characteristics, consisting of frequencies and percentages for categorical values and mean and standard deviations (SD) for scale variables. For the mental health variables (anxiety and depression), skewness and kurtosis values were obtained and the Shaprio-Wilk Test was performed to assess normal distribution (unimodal, skewness<1; Supplementary table 1). Bivariate associations between mental health variables and age (continuous variable) were assessed via Pearson’s correlation coefficient *r*. Significant differences in mean level of mental health variables between categories of dichotomous sociodemographic variables were assessed via t-test. The associations between mental health variables and sociodemographic variables were assessed in the following groups: by living status (alone vs not), previous psychiatric diagnosis (yes no), current intake of psychopharmacological drugs, high risk for COVID-due to comorbidities, and high frequency of social contact (>6 social contacts per week vs less than 6).

**Table 1.** Demographic and clinical characteristics assessing medical history, previous psychiatric and psychotherapy treatment and experience. Total N represents the number of callers which answered the question. N represents the number of callers which affirmed the features asked. The percentage was calculated as (N/total N)*100.

|  | Total N | N | (%) |
| --- | --- | --- | --- |
| **Known co-morbidities** | 52 | 46 | 88,5 |
| Cardiovascular | 52 | 24 | 46,2 |
| Chronic lung disease | 52 | 14 | 26,9 |
| Diabetes | 52 | 8 | 15,4 |
| Current or previously diagnosed cancer | 52 | 12 | 23,1 |
| Neurological diseases | 52 | 10 | 19,2 |
| **Number of daily taken medications** |  |  |  |
| </=1 | 48 | 13 | 27,1 |
| >1 | 48 | 35 | 72,9 |
| **Kind of daily medication** |  |  |  |
| Psychopharmaca | 48 | 15 | 31,3 |
| Antihypertensiva or cardial medication | 48 | 18 | 37,5 |
| Anti-diabetic medication | 48 | 6 | 12,5 |
| COPD or asthma medication | 48 | 10 | 20,8 |
| **Pre-diagnosed with psychiatric disease** | 52 | 20 | 38,5 |
| Anxiety | 52 | 4 | 7,7 |
| Depression | 52 | 17 | 32,7 |
| Psychosis | 52 | 1 | 1,9 |
| Addiction | 52 | 4 | 7,7 |
| Currently seeing a psychiatrist | 52 | 12 | 21,8 |
| Previously seeing a psychiatrist | 52 | 5 | 9,1 |
| **Psychiatric treatment** |  |  |  |
| Currently seeing a psychiatrist | 52 | 11 | 21,2 |
| Previously seeing a psychiatrist | 52 | 5 | 9,6 |
| Never in psychiatric treatment | 52 | 33 | 67,3 |
| **Previous Psychotherapy experience** |  |  |  |
| No Previous Therapy | 52 | 35 | 67,3 |
| Consultation or less than 6 Months | 52 | 7 | 13,5 |
| Psychotherapy more than 6 months | 52 | 4 | 7,7 |
| Continuous Psychotherapy over years | 52 | 6 | 11,5 |

#### Ethics approval and consent to participate

Informed consent was obtained from the subjects when calling our hotline to use data collected during the call for scientific purposes according to the statues of the ethics commission of the Faculty of Behavioral and Cultural studies at the University of Heidelberg. Written consent could not be received as this was an anonymous hotline. We have presented the study design and data acquisition to members of the ethics committee II at the medical faculty Mannheim, University of Heidelberg. Ethical approval was waived due to the anonymous design of the hotline.

## Results

A total of 55 older adults called the COVID-19 Psychiatric hotline for older adults during the initial stage of the pandemic (April 13- June 15). We received a higher frequency of numbers of calls during the initial peak of the COVID-19 pandemic in Germany in April 2020 compared to end of May and June, 2020 (Fig 1A). The reasons for calling the hotline varied, with most reported reasons being: general information about corona (60.7%), requesting health and psychological problems (32.1%), psychiatric consultation (26.8%), loneliness (21.4%) or requesting help with problems of daily function (7.1 %) (Fig 1B). The average duration of each call was 39.56 mins (SD = 22.93, range 9-120 mins).

**Figure 1.**
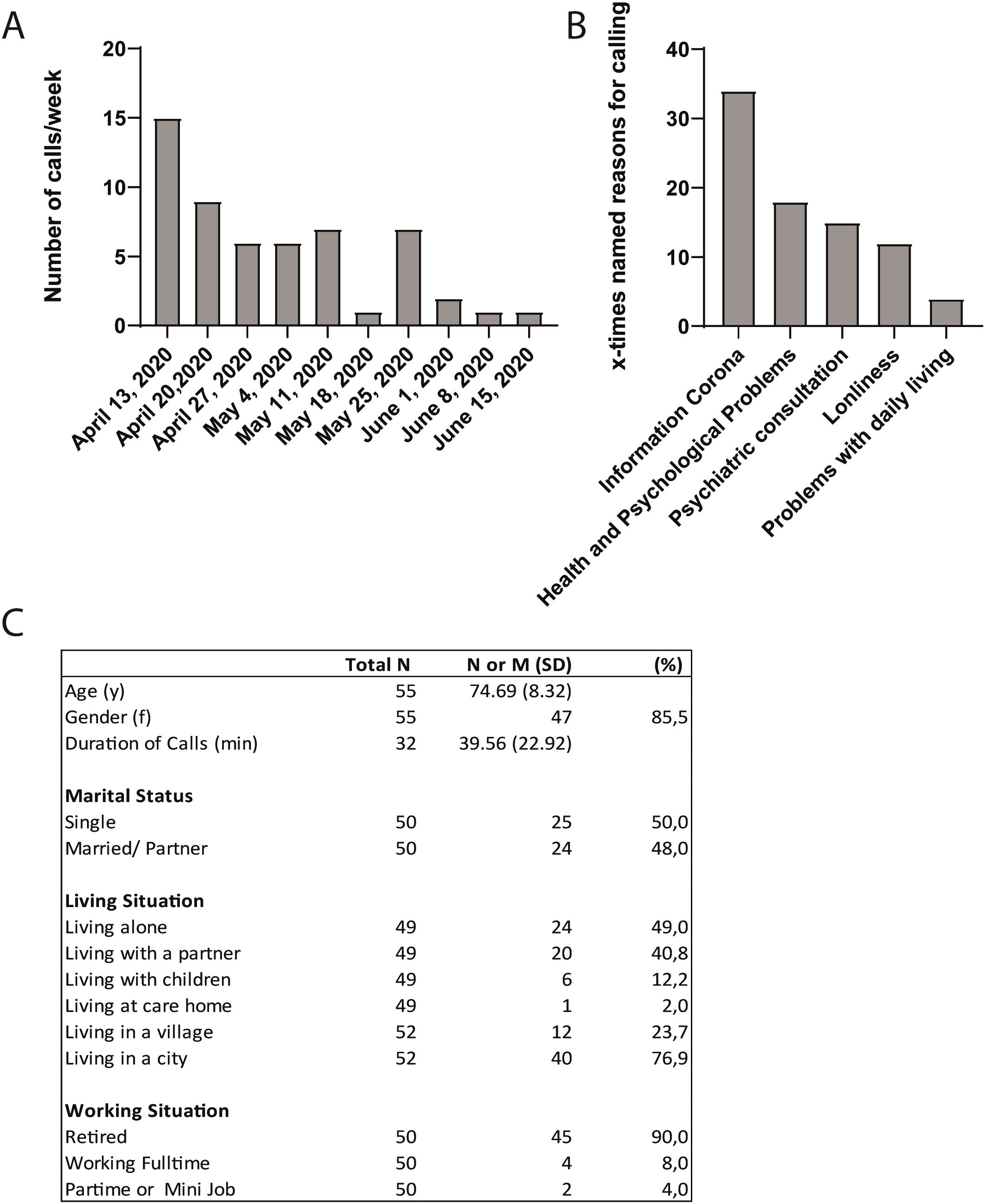
(A) Histogram showing the frequency of calls/week received at the geriatric helpline. (B) Graph depicting the reasons for calling the geriatric hotline. (C) Demographic characteristics of callers. Total N represents the number of callers which participated which answered the question. N represents the number of callers which affirmed the feature asked. The percentage was calculated as (N/total N)*100.

### I Demographics

53 respondents completed the full survey. Sociodemographic characteristics are presented in Fig.1C. Most callers were women (85.5%), older adults (Age *M* = 74.69 SD = 8.32; range 59-98), single (50.0%) living in a city (76.9%) and retired (90.0%). Almost half or the callers were living alone (49.0%), with the rest either living with their spouse or partner (40.8 %) or with their children (12.2%) and one caller was living in a care home.

#### Medical History

46 (88%) callers reported known co-morbidities, with cardiovascular, (N=24, 35%) chronic lung disease, (N=14, 20%) Diabetes, (N=8, 12%) current of previously diagnosis of cancer (N=12 18%) and neurological diseases (N=10, 15%) among the most common comorbidities (Table 1). The average Charlson Comborbidity Index (CCI) score was 4.02 (SD = 1.52; range 0-11). 45% of respondents (N =23) were considered at high risk for COVID-19 based on their comorbidities of either COPD, diabetes and cardiovascular disease. In terms of medication intake, 37% were prescribed with antihypertensive or heart drugs versus 12% of diabetes or COPD 20% medication. Most callers (72%) took more than one medication daily (Table 1).

#### Psychiatric and psychotherapy experience

38.5% of the callers were pre-diagnosed with a psychiatric disease, out of which, depression (32%) and anxiety disorders (7.7%) were the most common diagnosis. Out of 52 respondents, 21% were currently seeing a psychiatric for treatment and 31% of callers received psychotropic drugs (Table 1). The majority of callers had no previous psychotherapy experience (67.3%) and 11.5% of callers had had continuous psychotherapy over years (Table 1).

### II Changes in daily life and social interaction

As expected, 52.8% of callers reported a decrease in their social contact since the COVID-19 crisis started, with 56.6 % having less conversations with contacts and 79.2% reported less visits from others, while only 5.7% of the callers reported an unclear support of daily living due to the COVID-19 crisis (Figure 2A).

**Figure 2.**
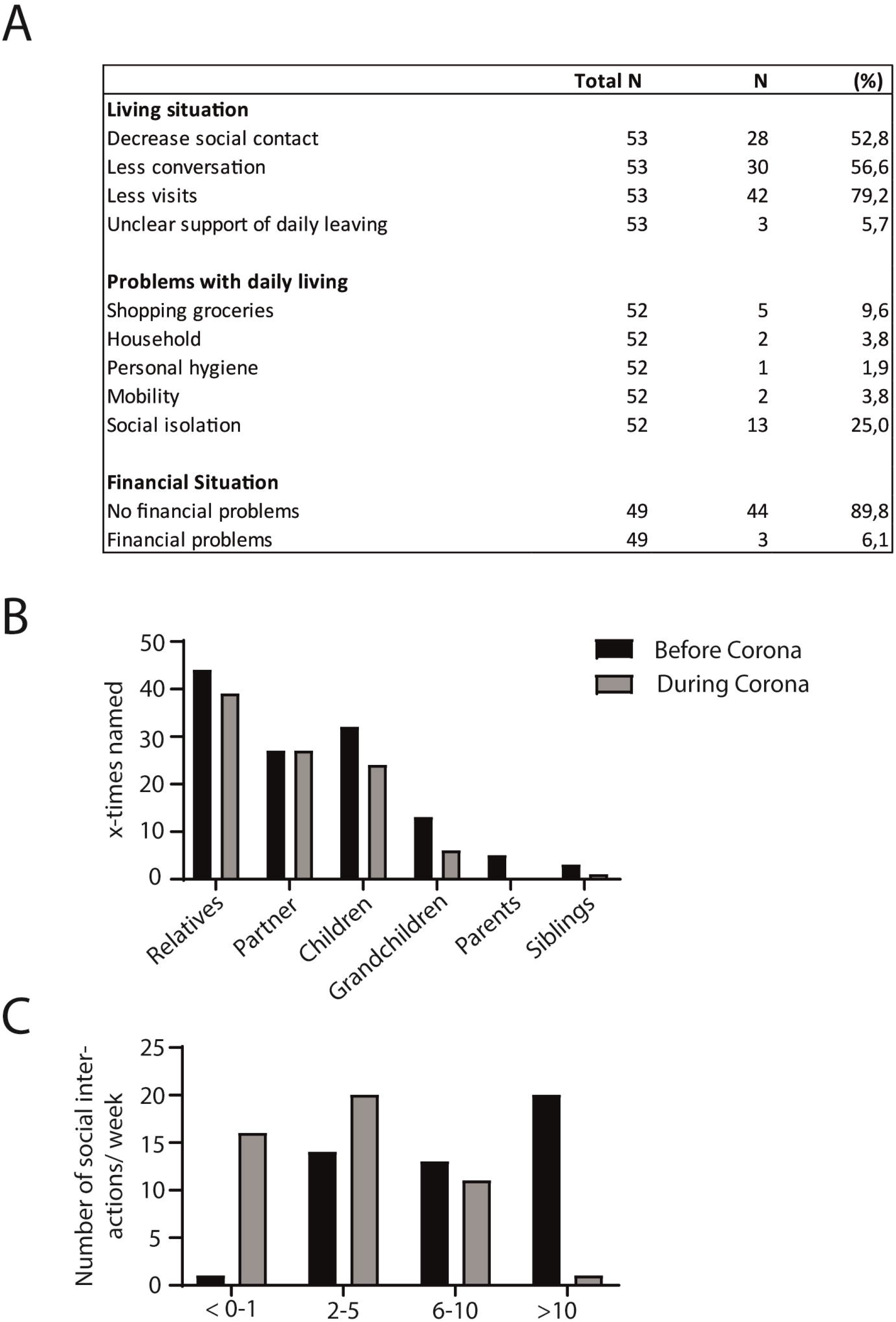
(A) Table depicting changes in daily living and in the financial situation due to the COVID-19 pandemic and lockdown measures. (B) While the social interaction to the partner remained stable, callers reported less social interactions with all other relatives (including children, grandchildren, parents and siblings) during the crisis. (C) The number of social interactions also decreased during the COVID-19 pandemic, while most participants had >10 social interactions in person per week before COVID-19, most callers reported a decrease of all personal social interactions of 2-5/week during the lockdown measures.

#### Significant Changes in daily life

When asked about their significant changes in daily life, 25% reported social isolation as a current problem in their daily life, while 9.6% reported problems accessing or shopping for groceries. Only one or two callers announced issues with personal hygiene or mobility (Figure 2A). New financial difficulties due to the Covid-19 crisis did not arise for most of the callers (89.8 %, Figure 2A).

#### Numbers and kind of social interactions change

Although interactions with the spouse and partners did not change because of social restriction in the initial peak of the Covid-19 pandemic, callers reported to have less direct, personal contact with all other relatives such as children, grandchildren, parents or siblings (Figure 2B). Before the Corona crisis 42% of callers had over 10 social interactions per week, which during the lockdown decreased to 1.8%. During the lockdown, the most common of number interactions per week was reduced to 2-5 per week (42.6%) and 0-1 (31%) (Figure 2C).

### III New Psychiatric Symptoms due to COVID-19

Over 69% of callers reported new or an increase in psychiatric symptoms during this time (Figure 3A) with anxiety (47.2%), depressed mood (56.%), sleep disturbances (30.2%), anhedonia (32.1%), restlessness/agitation (22.6%), changes in cognition (concentration difficulties) (15.4%) and suicidal thoughts (15.1%) among the most common reported symptoms.

**Figure 3.**
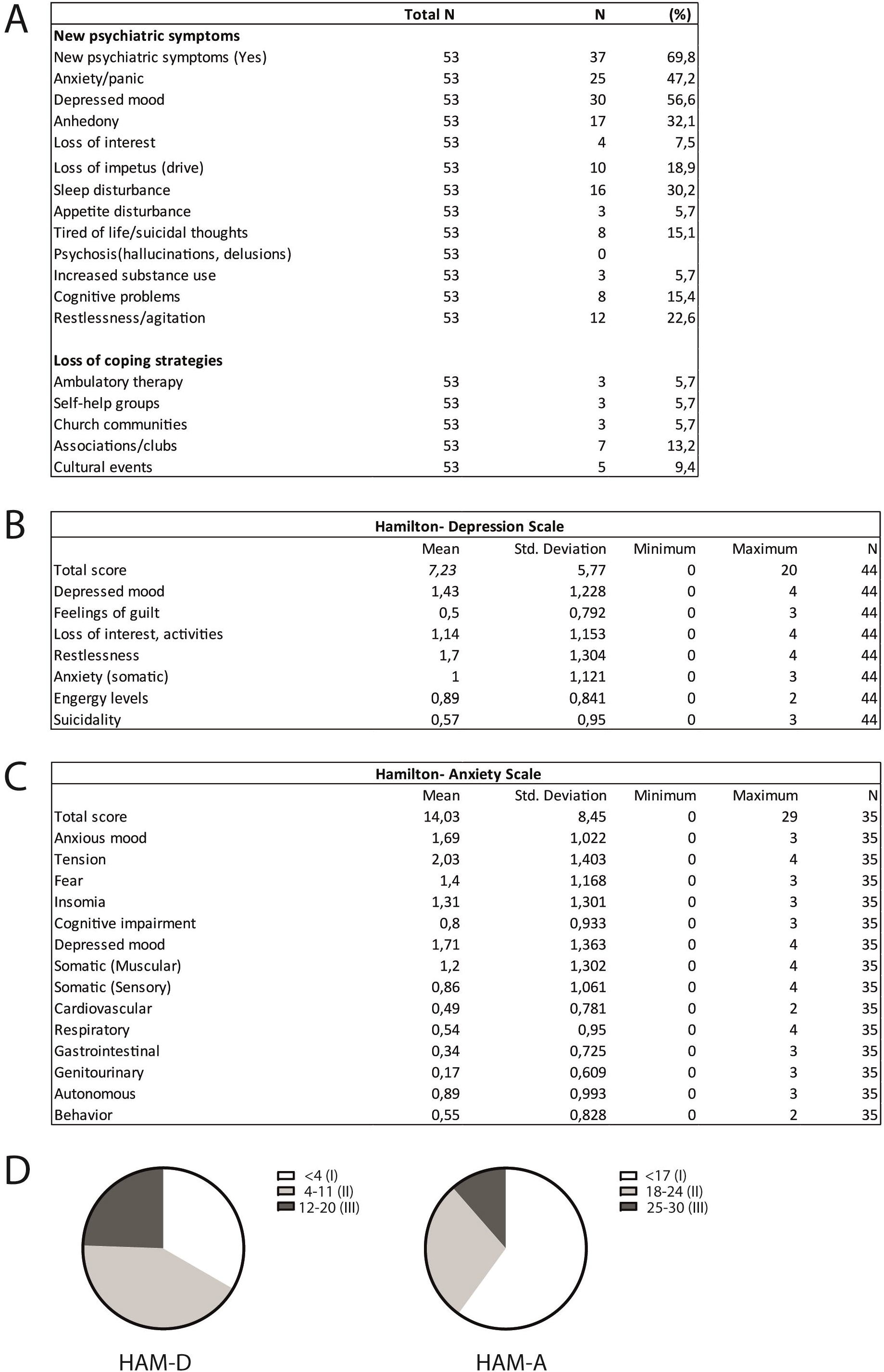
(A) Reported new psychiatric symptoms and loss of coping strategies during the Covid-19 pandemic. N represents the number of callers which affirmed the different features asked. The percentage was calculated as (N/total N)*100. (B) Mean scores and frequency of the - Hamilton Depression Rating Scale (HAM-D) and (C) the Hamilton Anxiety Rating Scale (HAM-A) assessments. (D) Pie charts revealing the level of depressive or anxiety symptoms: A third (33.3%) showed no depressive symptoms (I), versus 42.2% with mild levels (II), versus 24.4% with moderate levels (III). For the HAM-A 60% of callers showed mild levels of anxiety (level I) versus 28.6% with moderate (II) and 11.4% with moderate to severe (III) levels of anxiety. We neither found severe (level IV) depressive nor anxious symptoms among the participants.

#### Loss of therapeutic resources

Around 5 % of callers reported loss access to therapeutic resources (ambulatory psychotherapy, self-help group and church meetings), while 13.2% of callers reported a loss of weekly activities through sports clubs or cultural associations (Figure 3A).

#### Mental Health Status

##### Depression

Symptomatic of depression, as assessed by the HAM-D7 score showed a mean score of 7.23 (SD = 5.77; range 0-20, Figure 3B, Supplementary Table 1). Regarding the subscales, the most reported symptoms were restlessness (M= 1.7), depressed mood (M =1.45) and anhedonia or loss of interest (M=1.14, Figure 3B). A total of 19 (42.2 %) were considered to have mild levels of depressive symptoms (Score 4-11); 11 (24.4%) showed moderate levels (score 12-20) while we did not find participants with severe depressive symptoms (score >20 according to (McIntyre et al., 2002), Figure 3D). A third of participants did not report depressive symptoms (score <4, Figure 3D).

##### Anxiety

A total of 35 participants completed the anxiety scale measured by the HAM-A and showed a mean score of 14.03 (SD =8.45; range 0-29, Figure 3C, Supplementary Table 1). The most reported symptoms were tension, depressed mood, anxious mood, fear, insomnia, and somatic muscular symptoms. Assessing the different anxiety levels as described (Hamilton, 1959), 60.0% of callers were considered to have mild levels of anxiety, (score <17, Figure 3D); 28.6% (N=10, score= 18-24) showed moderate and 11.4% (N=4, score= 25-30) moderate to severe levels of anxiety. No participants revealed severe anxiety symptoms (score>30, Figure 3D).

### III Associations between demographics variables, social changes and mental health status during the pandemic

Age was significantly negatively correlated to higher levels of anxiety (*r* = -.340 p =.045) and depression symptoms (*r* = -.293 p =.054, Table 2), showing that older individuals report less symptoms. Individuals pre-diagnosed with a psychiatric disease reported significantly higher levels of depressive (*t*(42) = 3.33, *p* = .001) and anxiety symptoms (*t*(33) = 4.00, *p* > 0.001) than those without a diagnosis. Likewise, those individuals who were currently taking psychopharmaceutical medication also reported significantly higher depressive (*t*(39) = 3.33, *p* =.002) and anxiety symptoms (*t*(31) = 2.97, *p* = .006). Individuals that lived alone reported significantly lower levels of symptoms of anxiety (*t*(33) = -.2.57, p = .015), with no differences reported for depression symptoms (*p* = .017, Table 2). There were no significant differences in depression or anxiety symptoms for individuals who were either considered at high risk for COVID-19 due to high co-morbidities, nor those engaging in frequent social contact.

**Table 2.**
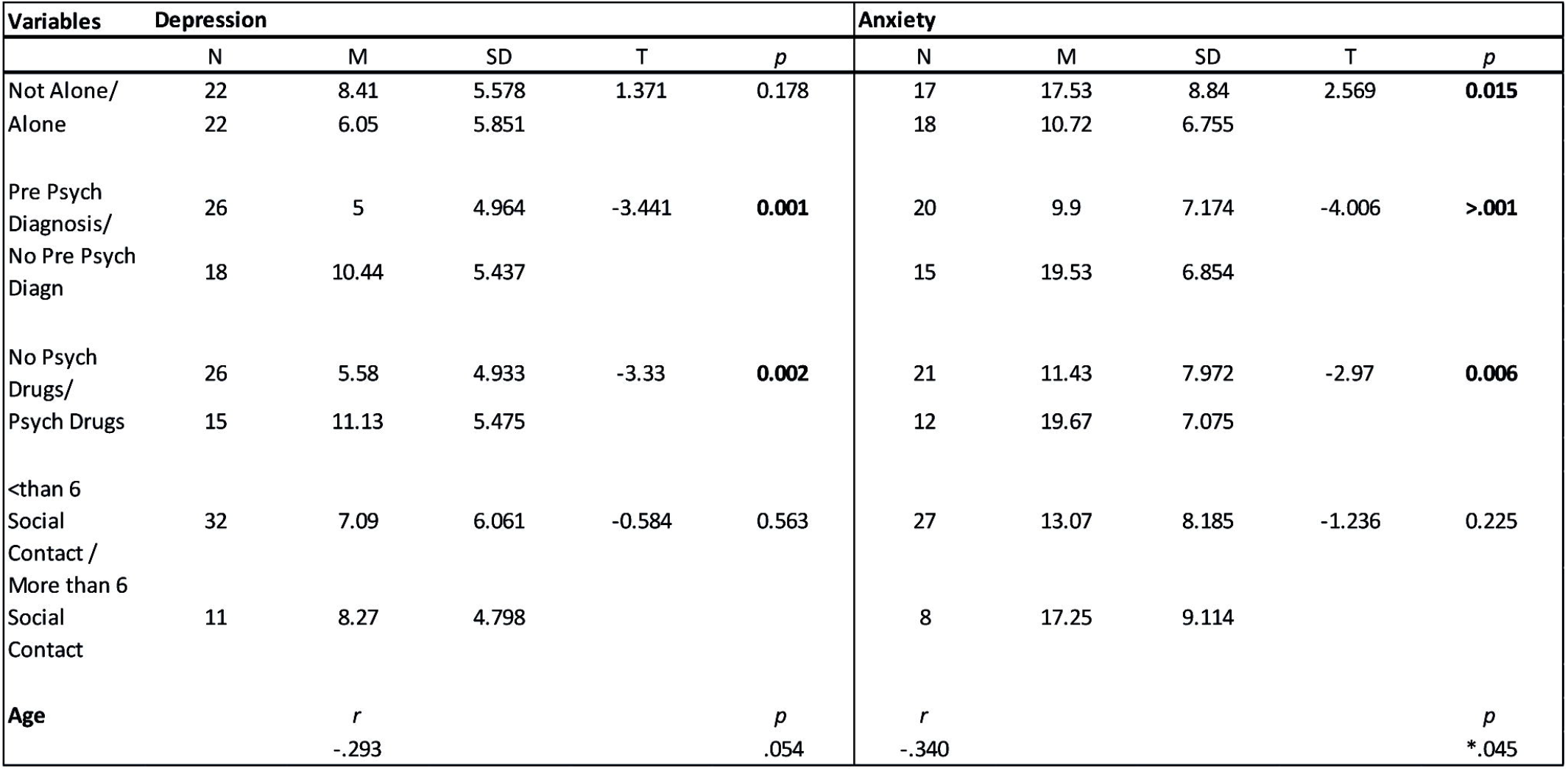
Associations between sociodemographic variables and mental health status. Differences in mean levels between categories were assessed via t-test. Individuals living alone reported higher anxiety symptoms than those living with others. Individuals with a pre-diagnosed psychiatric disorder reported more anxiety and depressive symptoms than callers without. Callers currently taking psychotrophic drugs reported significantly more depressive and anxiety symptoms. There were no difference among those at higher risk for COVID-19 or those engaging in frequent social interactions. Age was negatively correlated with higher levels of depressive and anxiety symptoms.

#### Help Provided

For almost two third of the callers (58.8%) we provided information of Covid-19 and recommended measurements to protect one-self (Supplementary Table 1). In 7 cases (13.7%) we were also asked for our professional estimation of a somatic comorbidity and referred to a doctor with the suitable specialization. For 29.4% (N=15) we recommended psychiatric counseling. Most callers (56.9%) required on open ear for the current situation and sorrows and reported of feeling significant relieved because of our psychological support. In rare cases we also provided practical help for daily living, social contacts and contact to welfare work (Supplementary Table 2).

In three quarters (74.5%) of all calls no subsequent treatment was required. However, for three participants (5.9%) we provided further psychiatric counseling in form of further psychiatric diagnostics and treatment. 7 callers (13.7%) received repetitive psychotherapeutic support with up to 6 subsequent telephone appointments. In one case (1.9%) we also called in our social worker (Supplementary Table 2).

## Discussion

As governments grapple with combatting the COVID-19 pandemic by implementing renewed restriction measures, our study elucidates the needs of older adults and the psychological impact during the initial stages of the health crisis. The long-lasting psychological consequences from this pandemic are largely unknown. However, our results from the geriatric helpline, help fill the gap by providing insight in to the timely efforts which mental health institutions can implement to cater specifically to older adults.

Our results showed that most callers where in need of general information about COVID measures, however a significant portion required help with psychosocial problems such as loneliness, daily functioning, and new psychiatric symptoms namely anxiety and depressed mood. The length of the calls implies the resources and level of involvement needed per caller. The frequency of the calls at the beginning, and after changes in the restriction measures, highlight the unclarity of the restriction measures and the need for further clarification from the authorities.

Our helpline was able to rapidly respond to the specialized psychosocial needs during the crisis. Our specified support, in particular psychiatric counseling and therapeutic talks, highlight the urgency to ameliorate the mental health impact of the COVID-19 in real time among older adults. One third of callers were recommend psychiatric counseling, while 13% of callers were in need and given continuous psychotherapeutic support. In a time where most mental health centers were closed and ambulatory therapy reduced their activity due to the lockdown measures, our results corroborates the need for the quick adaptation of mental health services and institutions. Ensuring continuity of psychiatric support and easy access to geriatric mental health care in the absence of standard practices are essential to mitigating negative mental health outcomes.

Older age is considered an important risk factor for COVID-19, where older adults are disproportionately negatively impacted. However, our results reveal that the psychological impact appears to ameliorate with age, with older adults reporting less symptoms of anxiety and depression. This is consistent with recent results and previous literature which identifies older age as a protective factor in dealing with disasters or crisis (Klaiber et al., 2020; Rodríguez-Rey et al. 2020). In the context of COVID, older adults showed better emotional well-being and were less reactive to COVID stressors than younger adults (Klaiber et al., 2020). Similarly, an initial study in China revealed that young individuals were at higher risk of suffering from anxiety than older adults during the outbreak (Huang & Zhao, 2020). In line with disaster crisis literature, older victims previously show lower anxiety, stress, and depression symptoms than younger individuals(Ngo, 2001) with researchers attributing this to their greater life experience, crisis exposure or by having to face fewer life responsibilities and life experience (Luchetti et al., 2020). Further studies should explore longitudinally this protective effect of age and its consequences in the context of the COVID-19 pandemic.

The majority of our callers reported either new or an increase of psychiatric symptoms. This highlights the psychological impact of COVID-19 among older adults population and supports the need for targeted psychological intervention strategies for seniors. Our results show that those individuals pre-diagnosed with a psychiatric disorder and those taking psychotrophic medication, reported higher anxiety and depressive symptoms than those without. Our findings add to the growing body of emerging literature which indicates that individuals with current and or past psychiatric disorders may be particularly vulnerable to the negative psychological sequalae of the pandemic(Benke et al., 2020; Guessoum et al., 2020; Meng et al., 2020). In contrast, individuals that were considered at higher risk for COVID-19 due to their somatic comorbidities did not report significantly higher levels of anxiety or depressive symptoms. This finding suggests that unlike psychiatric comorbidities, somatic comorbidities may not play a significant role on mental health outcomes in older adults. However, this should be further studied in larger samples.

In accordance to other studies across Europe, most callers have experienced changes in their daily life, with social isolation being considered a problem in a quarter of callers (Benke et al., 2020; Rodríguez-Rey et al., 2020). Interestingly, our results show that living alone proved to be a protective factor against anxiety symptoms, as individuals cohabitating with others reported higher levels of anxiety. This is in contrast with previous findings from general population surveys, which find cohabitation to be a protective factor against psychological suffering and negative mental health outcomes (Benke et al., 2020; Rodríguez-Rey et al., 2020). Contrary to expectations, our data reveals that older individuals living alone were less anxious and thus coped better. Older individuals living alone were perhaps better able to cope with the social distancing rules or “stay-at home” orders since they were more used to living alone for longer. Given the scarce and mixed evidence to this date, this possible psychosocial resilience factor merits further attention, specifically in older adults.

The limitations of this study should be considered when interpreting the findings. First, this data was collected during the initial stages of the pandemic. Concerns, daily experiences and mental health outcome may change and evolve as the outbreak evolves. Second, our sample was limited, as advertised, to cater to older adults over the age of 65, therefore age-related associations and differences must be treated with caution. Finally, our sample may not generalize to other populations as it is entirely subject to individual’s motivation to call the helpline.

## Conclusion

The present study elucidated the specific needs of older individuals through the implementation of a geriatric helpline. The rapid adaptation of mental health institution’s resources through such initiatives are necessary to support psychosocial needs and psychological well-being of older adults during a health crisis. Considering the pandemic will most likely have lasting effect, ongoing help initiatives and follow-up studies are warranted to mitigate negative health outcomes in the vulnerable population.

## Data Availability

The authors declare that all data supporting the findings of this study are available within the paper and its supplementary material. Raw data can be made available upon reasonable request.

## Acknowledgements

Not applicable

## Tables and Figure legends

**Supplementary Table 1**. Skewness and Kurtorsis for mental health variables of depression and anxiety. SE Standard Error. Both variables were sufficiently normally distributed.

**Supplementary Table 2**. Table depicting the different features of help provided and subsequent appointments scheduled by us for psychiatric or psychotherapeutic counseling or support by our social worker. Total N represents the number of callers which participated in the survey. N represents the number of callers which affirmed the different features asked. The percentage was calculated as (N/total N)*100.

