## Supplemental Table 1 and 2 for "Rapid Support for Older Adults during the initial stages of the COVID-19 Pandemic: Results from a Geriatric Psychiatry helpline"

### Supplementary Material

**Supplementary Table 1.** Skewness and Kurtorsis for mental health variables of depression and anxiety. SE Standard Error. Both variables were sufficiently normally distributed.

**Supplementary Table 2.** Table depicting the different features of help provided and subsequent appointments scheduled by us for psychiatric or psychotherapeutic counseling or support by our social worker. Total N represents the number of callers which participated in the survey. N represents the number of callers which affirmed the different features asked. The percentage was calculated as (N/total N)*100.

| | | Skewness | | Kurtosis | |
|---|---|---|---|---|---|
| | **N** | **Value** | ***SE*** | **Value** | ***SE*** |
| Anxiety | 35 | -0.058 | 0.398 | -1.271 | 0.778 |
| Depression | 44 | 0.293 | 0.357 | -0.908 | 0.702 |

**Supplementary Table 1** SD= Standard deviation. SE= Standard error

| | Total N | N or M (SD) | (%) |
|---|---|---|---|
| **Result of the hotline call** | | | |
| Providing information of Covid-19 | 51 | 30 | 58.8 |
| Practical help/information for daily living | 51 | 1 | 1.9 |
| Providing social contacts | 51 | 1 | 1.9 |
| Providing help for somatic diseases/issues | 51 | 7 | 13.7 |
| Recommendation for psychiatric counseling | 51 | 15 | 29.4 |
| Providing relieving psychological support | 51 | 29 | 56.9 |
| Providing contact to welfare work | 51 | 1 | 1.9 |
| **Subsequent treatments** | | | |
| No subsequent treatments | 51 | 38 | 74.5 |
| Subsequent treatments required | 51 | 10 | 19.6 |
| Subsequent psychiatric treatment provided | 51 | 3 | 5.9 |
| Subsequent psychotherapy provided | 51 | 7 | 13.7 |
| Subsequent welfare work provided | 51 | 1 | 1.9 |

**Supplementary Table 2**
